# Public Service Quality under Permanent and Fixed-term Employment Contracts: Evidence from Physician Supply of Primary Care

**DOI:** 10.1101/2021.04.22.21255930

**Authors:** Sean Sylvia, Hongmei Yi, Hao Xue, Gordon Liu

**Affiliations:** University of North Carolina at Chapel Hill; Peking University; Stanford University February 2021

**Keywords:** Primary Health Care, fixed-term physicians, Headcount Quota System, Standardized Patients, China

## Abstract

A key feature of public sector employment in many countries is rigid civil service rules that effectively limit manager autonomy over hiring, firing, promotion, and compensation decisions. We study the effect of these rules by comparing the quality of healthcare provided by physicians employed as civil servants with physicians hired in the same facilities on fixed-term contracts that give managers more autonomy over personnel decisions. Using data from interactions with unannounced standardized patients, we find that fixed-term contracts motivate greater diagnostic effort without increasing unnecessary treatments. Lower effort among civil servants appears due to both weaker career and wage incentives.

## H1

A feature of public sector employment in many countries is rigid civil service regulations that govern hiring, firing, promotion, and compensation decisions (Finan et al., 2017). A primary consequence of these rules is often to constrain the autonomy of managers and others over these personnel decisions, effectively granting civil service employees with guaranteed long-term employment and compensation increases linked to tenure. Concern that this feature of civil service employment contributes to poor quality services has motivated reforms in several high and lower-income countries giving public sector managers greater autonomy over personnel decisions (Shim, 2001; Garćıa-Prado and Chawla, 2006; Bossert and Mitchell, 2011; Cotlear et al., 2015; Cobos Muñoz et al., 2017; Banzon and Mailfert, 2018). The impact of these reforms on service delivery, however, remains understudied empirically, as is the underlying hypothesis that rigid civil service rules reduce service quality (Dwicaksono and Fox, 2018; Finan et al., 2017; Rasul and Rogger, 2018). In particular, it remains unclear whether often-documented quality deficits in government-run facilities are due to civil-service employment contracts directly or to the nature of the public sector more generally.

This study aims to contribute evidence on this issue by comparing the performance of civil service physicians to those employed through fixed-term contracts in the same government-run facilities in China. Although most physicians are employed under the *bianzhi* system (China’s quota-based system regulating civil service posts), facilities have flexibility to hire fixed-term physicians “beyond quota” (Tao, 2014; Zhang et al., 2012). Those employed under the *bianzhi* system as civil servants (henceforth, “civil service physicians”) are deployed and remunerated by upper administrative units and are said in China to have an “iron rice bowl,” as they face a low probability of dismissal and are nearly virtually ensured long-term, if not lifelong, employment (Liu et al., 1996). In contrast, facility managers have autonomy over hiring and firing fixed-term physicians (“fixed-term physicians”) and are responsible for their salary and benefits (Eggleston et al., 2008; Ang, 2012; Zhang et al., 2012; Tao, 2014; Mao, 2017). Whether to reform the *bianzhi* system has in fact been the subject of substantial policy debate in the context of broader health reforms in China (World Bank, 2019). Some argue that the system prevents the efficient allocation of the health workforce while others argue that employing physicians on fixed-term contracts would lower health service quality (Yip and Hsiao, 2014; Xu, 2014; Ren and Wang, 2015; Tao, 2014; Wu, 2014). There is currently a marked lack of research on the *bianzhi* system to guide this debate (Wang, 2021).

In comparing physician performance under civil service and fixed-term contracts, we address potential sources of endogeneity by accounting for patient mix and the sorting of physicians across facilities. We use data from an audit study in which standardized patients (SPs) were extensively trained to consistently present three disease cases—unstable angina, gastroenteritis, and pulmonary tuberculosis—and were sent unannounced to a random sample of 208 township health centers in three separate provinces. Using these interactions, we evaluate physicians on clinical process (adherence to a checklist of recommended questions and exams for each disease case), appropriateness of treatment, and costs. This audit methodology allows us to control for potential differences in patients across physicians because the disease cases presented are fixed by design (Das et al., 2012; Leonard and Ottar, 2016; Das et al., 2016b). To account for physician sorting, we further control for facility fixed effects to compare civil service and fixed-term physicians in the same facilities. Without controlling for physician characteristics, we estimate the “as is” difference between the two types of physicians, which includes differences attributable to training, experience, and other factors. We also estimate the effects of fixed-term versus civil service contracts on performance by controlling for physician characteristics, including a measure of clinical competence specific to the diseases presented by SPs as well as measures of physician intrinsic and prosocial motivation. Thus, although we lack a source of exogenous variation, our study design allows us to produce plausibly exogenous estimates of causal effects under unconfoundedness.

We present several key results. First, fixed-term physicians substantially outperform civil service physicians in the same facilities. They perform more than 0.5 standard deviations higher on a measure of clinical process quality, a difference that persists after controlling for observed physician characteristics, including disease specific clinical knowledge and measures of internal motivation. Second, although fixed-term physicians may face stronger incentives to increases clinic revenue, we find no evidence that they are more likely to prescribe unnecessary drugs or that patients’ out-of-pocket costs are higher when they are seen by a fixed-term physician. Third, differences in performance are due to differences in effort: civil service physicians exhibit significantly larger gaps between their disease-specific knowledge and practice in treating patients. Fourth, the effect of fixed-term contracts relative to civil service are larger among more prosocially-motivated physicians. Fifth, pay is significantly correlated with performance for fixed-term physicians but not for civil service physicians. This suggests that the superior performance of fixed-term physicians may be due in part to stronger wage incentives.

Our findings contribute to the growing literature on the “personnel economics of the state” (Finan et al., 2015). A major focus of this literature has been on the incentives surrounding the selection and motivation of public sector workers in developing countries. Several studies have analyzed the employment of fixed-term contract teachers in public schools, on balance finding that students of contract teachers perform at least as well as students of civil service teachers on standardized examinations despite civil service teachers’ having superior qualifications and higher pay.^1^ Along with the different setting of healthcare, there are two important distinctions with our setting. The first is that managers had limited control over hiring and remuneration in the contexts that these studies analyze.^2^ As a result, researchers have emphasized the use of contract teachers as a “task-shifting” cost-reduction policy. In our context, the distinction between civil service and fixed-term physicians is better understood as a difference in managerial control induced by somewhat arbitrary quotas in China’s *bianzhi* system. Second, there is different scope in schools for compensating responses to the hiring of contract employees to affect quality: contract teachers can be assigned to different tasks within schools that complement or substitute for the effort of existing teachers (Duflo et al., 2015; Muralidharan and Sundararaman, 2013). Our setting compares physicians who all perform the same task of treating outpatients, and there is little reason to believe (or evidence showing) that civil service physicians reduce effort in their treatment of individual patients in response to other physicians being employed on fixed-term contracts.^3^

A broader value of exploring this question in the context of healthcare can also be seen in light of the question of in-house provision versus contracting-out of public services. In a sense, employing front-line workers on fixed-term contracts in government-run facilities is an intermediate form of contracting that lies between public and private provision. Hart et al. (1997) argue that in-house provision of public services is stronger “when non-contractible cost reductions have large deleterious effects on quality,” while stronger case for contracting out “when quality-reducing cost reductions can be controlled through contract or competition.” In-house (government) provision of healthcare may therefore be preferable if consumers have difficulty assessing quality, thereby limiting the potential role of competition to reduce costs. Hart el al., however, explicitly note that analyzing alternative arrangements in healthcare is “a great deal more complicated” than the case explored in their model. In a study methodologically similar to this one, Das et al. (2016b) compare public and private providers in India, finding that private providers exert significantly more effort in the private sector and are not more likely to prescribe unnecessary treatments. To interpret their findings, they present a model illustrating how the ultimate effect on overall quality likely depends on how the marginal gains in diagnostic effort from market incentives compare to increased costs due to over-treatment. Here, our findings imply that providing facility managers greater control over physician employment may outperform public or private provision if quality and costs are more observable to managers than consumers, and provided that managers have appropriately-aligned incentives.^4^

How contract types affect over-treatment and interact with internal motivation are also particularly relevant in the public sector because publicly-provided services often have multiple, ill-defined goals and prosocial motivation is an important driver of selection into public service (Dixit, 2002; Besley and Ghatak, 2005).^5^ In this context, there is concern that approaches to increase effort among civil servants using extrinsic rewards such as performance pay may distort effort away from less observable tasks or could ‘crowd-out’ prosocial motivation (Holmstrom and Milgrom, 1991; Bénabou and Tirole, 2006; Dhaliwal and Hanna, 2017; Ashraf et al., 2019). It has also been noted that findings of lackluster effects of performance pay in some settings might be, in part, because performance pay schemes have not generated incentives that are strong enough to counteract existing institutional incentives, such as those stemming from civil service regulations (Eijkenaar et al., 2013; Das et al., 2016a; Mendelson et al., 2017; Patel, 2018).^6^ Our results suggest that career concerns and inherent relational contracts may be a more fundamental determinant of quality and do not crowd-out the internal motivation of public servants.^7^

The remainder of the paper is organized as follows. In the next section, we briefly introduce the *bianzhi* system and physician deployment practices in rural China. In Section 3, we discuss the details of data collection and empirical approach. Section 4 presents the results, and we discuss implications and conclude in Section 5.

## 2 The *Bianzhi* System and Physician Deployment in China

Since the establishment of the People’s Republic of China in 1949, the public healthcare system and other public institutions have been staffed through a head-count quota system for civil service posts referred to as *shiye bianzhi* (SCOPSR, 2010a,b). The *bianzhi* system defines the total number of personnel to be employed in public facilities and the composition of each facility’s workforce in terms of posts, grades, and professional titles. Approved bianzhi posts are funded by the finance bureau, and those employed under *bianzhi* have civil service positions with their public service unit. Public physicians in rural areas are generally employed by county health bureaus. Those employed in civil service positions under *bianzhi* receive a fixed salary and may be eligible for bonuses and a variety of fringe benefits, including pensions, housing subsidies, and insurance (SCOPSR, 2010a; Xu, 2014). Civil service employee performance is evaluated in yearly reviews under the “cadre evaluation system” (*kaohe*), yet in practice promotion decisions are dominated by seniority rather than performance. In common with many other countries, civil servants in China face little possibility of dismissal (although they may be reassigned). Although the concept originally referred to a broader set of economic policies, “iron rice bowl” is still used today to refer to employment under *bianzhi* due to the virtual guarantee of lifetime employment with a stable salary and benefits (Mao, 2017; Wang and Xing, 2016).

Historically, health facility managers have had little flexibility to hire staff outside of the *bianzhi* system. Prior to market-oriented reforms in the late 1970s and early 1980s, staff were universally supported by the nationally run Cooperative Medical Scheme (CMS). Following economic reforms and the resulting collapse of the CMS, fiscal responsibility was transferred to local governments; however, grassroots facilities had little incentive or means to hire staff beyond their *bianzhi* allocation due to a combination of declining utilization and rising input prices (Si, 1987; CNHEI, 2005; Liu et al., 1996; Liu and Yi, 2004; Ministry of Health of China, 2004; Yan et al., 2005). This situation changed with the initiation of demand-side health reforms around 2003. With increased demand, revenue, and availability of health insurance, local facilities began to employ large numbers of fixed-term physicians “beyond-quota” (Babiarz et al., 2012; SCOC, 2009; Wagstaff et al., 2009; Ren and Wang, 2015). At the same time, the government issued more rigid civil service regulations (Lu, 2003; SCOPSR, 2011).^8^ Facilities in rural areas found it difficult to recruit qualified personnel under heightened criteria for employment under civil service positions and began to rely on fixed-term physicians to meet growing demand (Ren and Wang, 2015). A nationally representative survey of six provinces, for instance, showed that, as of 2013, 22 percent of personnel at township health centers (the focus of our study) were beyond-quota (Ren and Wang, 2015). Other regional and case studies suggest a higher share (30-50 percent) at township hospitals in some areas (Ministry of Health of China, 2012; Zhang et al., 2012; Tao, 2014; Wei et al., 2015).

How physicians should be employed and remunerated is a significant issue in the context of broader reforms of the health system that are taking place in China. A key aim of recent reforms has been to strengthen primary care, including payment reforms designed to better align physician incentives with social objectives (Yip et al., 2012; Liu et al., 2017; Yip et al., 2019). With the advance of these reforms, the *bianzhi* system has been the subject of growing public discussion. Some have argued that the system restricts the efficient allocation of the health workforce as benefits are tied to specific units and that it constrains the ability of health facility managers to manage employees and to recruit staff according to facility needs (Yip and Hsiao, 2014; Xu, 2014; Ren and Wang, 2015; World Bank, 2019). The tying of budget allocations to the quota system also means that subsidies to facilities are largely unrelated to performance (Li, 2016; Mao, 2017). Opponents of *bianzhi* reform, however, argue that employing physicians on fixed-term contracts could lower health service quality (Tao, 2014; Wu, 2014) and that the need to hire off-bianzhi could, absent broader financing reforms, accelerate increasing costs by increasing pressure on health facilities to generate revenue (Xu, 2014). Despite this policy debate, there is currently no objective evidence on the relative performance of civil service physicians and those employed on fixed-term contracts.

## 3 Data and Empirical Approach

### 3.1 Sampling of facilities

The sample of township health centers used in this study was drawn to be representative of three prefecture-level cities (the administrative unit below the province) in Shaanxi, Sichuan, and Anhui provinces. One prefecture from each province was chosen for having a predominantly rural population and in consultation with local authorities. Across three selected prefectures, we randomly sampled 21 counties (out of 24 total counties across the three prefectures). Ten health centers were randomly chosen in each county, excluding the health center of the urban township in each county. Because one county had only nine eligible townships, the final sample consists of 209 township health centers (out of the 311 total existing in the three prefectures).

### 3.2 Data Collection and Summary Statistics

We conducted three separate waves of data collection in the sampled facilities. An initial facility and physician survey was conducted in June 2015. Five weeks later (August 2015), we measured the physician quality by collecting the data during unannounced visits by SPs to sample physicians. In September 2015, we tested physicians using clinical vignettes for the same cases presented by the SPs.

#### 3.2.1 Facility and Physician Survey

In each health center, a facility survey was administered to the facility director. This survey collected information on staffing and employment practices, availability of medical equipment and drugs, and characteristics of the population served by the clinic. The specific facility-level variables used in our current analysis include the number of civil service and fixed-term physicians employed, population in the catchment area of the township, total value of medical equipment in the facility, number of village clinics managed by the health center, total clinic revenue in the prior fiscal year, and distance to the nearest county-level hospital (the next-highest tier of the rural health system).

In each facility, we also interviewed all physicians who provided primary care. The physician survey gathered detailed information on physician demographics, employment contracts, experience, general and medical education, and physician certification level. We also collected information on prosocial and intrinsic motivation, using psychological scales developed by Grant (2008). The prosocial scale reflects motivation to “expend effort to benefit others,” and the intrinsic scale reflects motivation from a “desire to expend effort based on interest in and enjoyment of the work itself.” We extract factor scores (predicted values of latent variables) from each scale separately and normalize each using the mean and standard deviation in the sample

The characteristics of civil service and fixed-term physicians in our sample are shown in Table 1. Fixed-term physicians comprise approximately 19% of the primary care physicians in our sample. Compared to civil service physicians, fixed-term physicians have significantly lower levels of medical training and medical certification. Notably, vignette scores that measure physician competence (described below) as well as prosocial motivation and intrinsic motivation scores do not differ significantly between the two physician types.

**Table 1.**
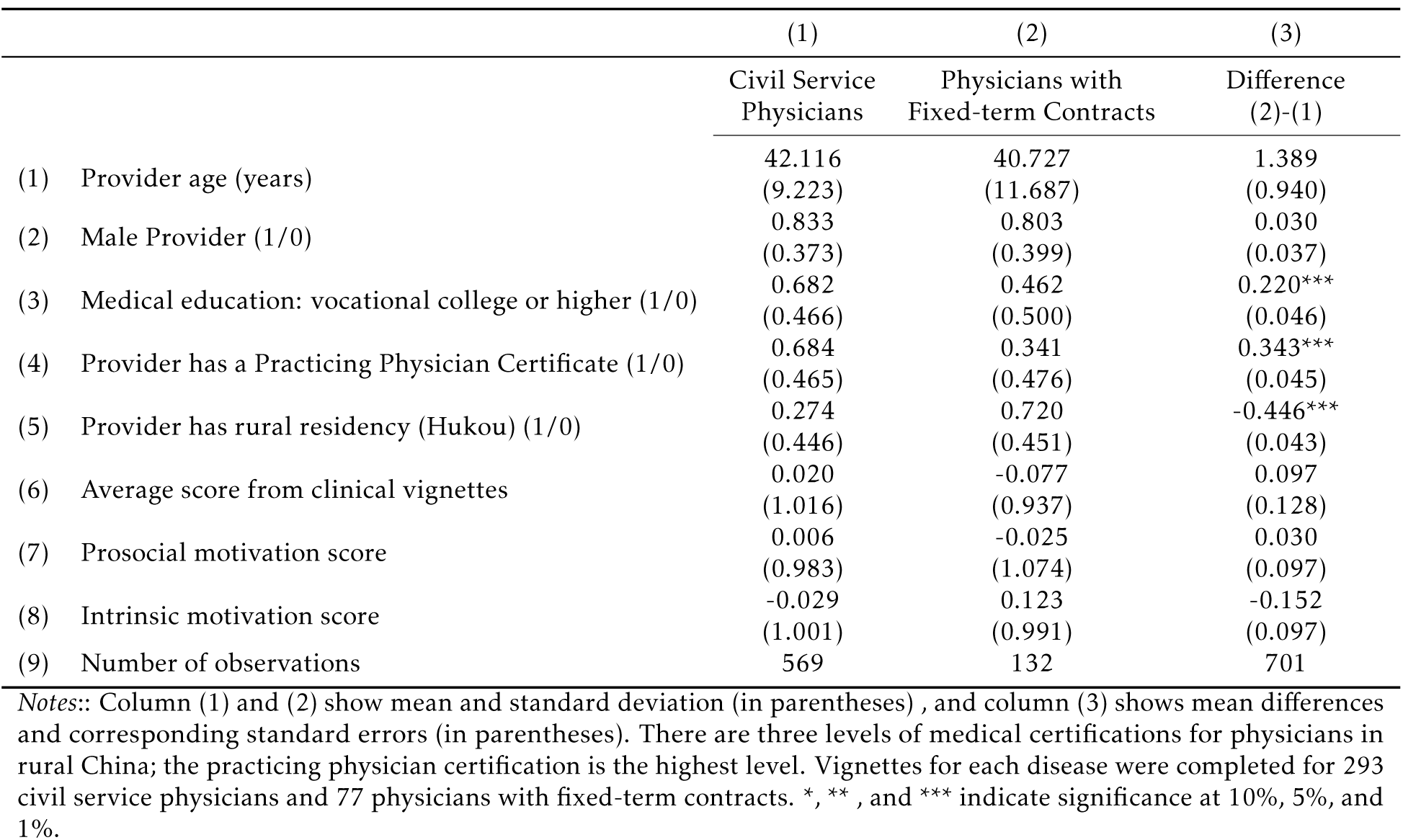
Physician Characteristics

Table 2 compares experience, pay and workload between physicians on the different contracts. Although civil service physicians have only 1.7 more years of experience, they have worked at their current facility for 3.6 years longer (12.2 vs. 8.7 years). Total pay (inclusive of wages and bonuses) is around 8% lower among fixed-term physicians. On average, civil service physician report that 35.6% of their pay was “performance-based” compared to 26.7% among physicians on fixed-term contracts. This amount was asked specifically in reference to bonuses explicitly designated as performance pay (referred to as “*jixiao gongzi*”). Though how this is determined varies, it often includes a portion tied to facility revenues and a portion tied to workload (Yip et al., 2010).^9^ In terms of workload, physicians on fixed-term contracts report working an additional 0.7 days per month, around 5 more hours per week, and seeing nearly 7 additional patients per week. Finally, 32% of current civil service physicians report starting their career on fixed-term contracts while over 90% of those currently employed fixed-term did so. Although we lack longitudinal data needed to explore the rate at which fixed-term physicians enter civil service, around 59% of current civil service physicians with 5-10 years of experience, 47% with 10-15 years of experience, and 62% of those with more than 15 years report starting as fixed-term.^10^

**Table 2.**
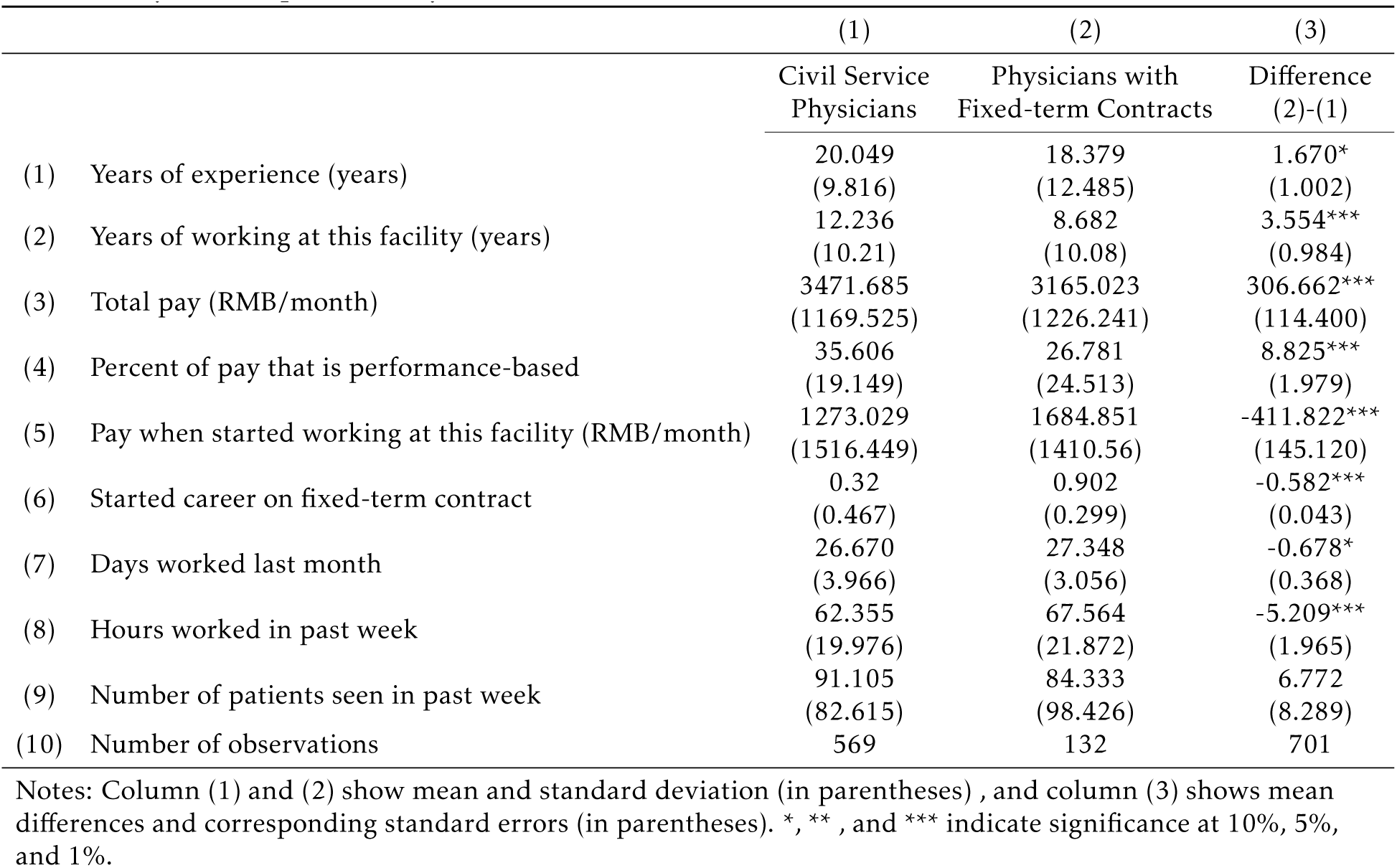
Physician Experience, Pay, and Workload

|  | (1) | (2) | (3) |
| --- | --- | --- | --- |
|  | Civil Service<br>Physicians | Physicians with<br>Fixed-term Contracts | Difference<br>(2)-(1) |
| (1) Years of experience (years) | 20.049<br>(9.816) | 18.379<br>(12.485) | 1.670*<br>(1.002) |
| (2) Years of working at this facility (years) | 12.236<br>(10.21) | 8.682<br>(10.08) | 3.554***<br>(0.984) |
| (3) Total pay (RMB/month) | 3471.685<br>(1169.525) | 3165.023<br>(1226.241) | 306.662***<br>(114.400) |
| (4) Percent of pay that is performance-based | 35.606<br>(19.149) | 26.781<br>(24.513) | 8.825***<br>(1.979) |
| (5) Pay when started working at this facility (RMB/month) | 1273.029<br>(1516.449) | 1684.851<br>(1410.56) | -411.822***<br>(145.120) |
| (6) Started career on fixed-term contract | 0.32<br>(0.467) | 0.902<br>(0.299) | -0.582***<br>(0.043) |
| (7) Days worked last month | 26.670<br>(3.966) | 27.348<br>(3.056) | -0.678*<br>(0.368) |
| (8) Hours worked in past week | 62.355<br>(19.976) | 67.564<br>(21.872) | -5.209***<br>(1.965) |
| (9) Number of patients seen in past week | 91.105<br>(82.615) | 84.333<br>(98.426) | 6.772<br>(8.289) |
| (10) Number of observations | 569 | 132 | 701 |

Notes: Column (1) and (2) show mean and standard deviation (in parentheses) , and column (3) shows mean differences and corresponding standard errors (in parentheses). \*, \*\*, and \*\*\* indicate significance at 10%, 5%, and 1%.

Summary statistics for facilities with and without fixed-term physicians are shown in Appendix Table 3. Around 38% of sample facilities employed fixed-term physicians. These facilities served a significantly smaller patient population. Although differences are insignificant, these clinics also tended have fewer civil service posts, to be slightly more remote, and to report a higher net revenue. In a regression predicting whether fixed-term physicians are employed these characteristics appear to explain little of the variation in the sample (Adjusted *R*^2^ = 0.07). The number of patients in the catchment area remains a strong predictor with an increase of 1000 patients correlating with a 0.8 percentage point decrease.

#### 3.2.2 Standardized Patients

Our primary measures of physician performance come from interactions between physicians and unannounced SPs. SPs have long been used as a pedagogical tool in medical schools and are an increasingly common tool for researching the quality of primary care (Madden et al., 1997; Das et al., 2012; Leonard and Ottar, 2016). A total of 63 SPs were recruited from local areas and intensively trained over a two-week period to consistently present disease cases to physicians. Each SP was trained to present either a case of unstable angina, a presumptive case of tuberculosis, or a case of child diarrhea (viral gastroenteritis). In the diarrhea case, SPs presented as the parent of an ill child. In each of these cases, SPs would engage physicians with an opening statement. Upon appropriate questioning by physicians, SPs would reveal symptoms consistent with the disease case that they were portraying. One SP presenting each disease case visited each facility, following the same procedures as any walk-in patient. Following clinical consultations with physicians, SPs purchased all medications prescribed and paid physicians their usual fee.

SP scripts from earlier studies in India (Das et al., 2012, 2015) were adapted to the Chinese context in collaboration with experts from medical schools in China and the China Centers for Disease Control and Prevention (the China CDC). In addition to an opening statement and set responses to potential questions by physicians, these scripts also contain detailed background information for each SP, including demographic characteristics, family situation, and occupation.

After each visit, SPs were debriefed using a structured questionnaire, and SP responses were confirmed against a recording of the interaction taken, using a concealed recording device. Data from SP interactions with physicians were used to construct several measures of care quality. The first metric is the adherence to a checklist of recommended questions and examinations to be completed by physicians. The checklist for each disease case was based on national and international clinical standards. To better discriminate between low-and high-quality physicians, we use completion of these checklist items to generate a composite quality index for each clinical interaction by estimating an item response theory (IRT) third-party logistics (3PL) model, following Das and Hammer (2005). Both the percentage of checklist items completed and IRT-scaled score are reported in our analysis.

In addition to this measure of process quality, we also construct outcomes based on whether the treatment/management of the patient is correct, total out-of-pocket costs incurred by the SPs, referral to upper-level facilities, the number of medicines prescribed, and the prescription of unnecessary or potentially harmful medication. The standards for correct treatment are shown in Appendix Table 4. Referral, although it may or may not reflect correct management, depending on the disease case, is included separately, given the important role of township health centers in the referral system. Referral may unnecessarily increase costs for certain cases (such as uncomplicated diarrhea) but is necessary for patients with symptoms of unstable angina or tuberculosis.

#### 3.2.3 Clinical Vignettes

Clinical vignettes were administered to each physician in a third round of data collection, following SP visits. These vignettes presented cases to physicians that were identical to those presented in SP visits. Vignettes were administered by two enumerators, one who played the role of the patient and the other who provided instructions and recorded the interaction. At the start, physicians were asked to proceed as they would with a real patient and were told that the patient would answer any questions and comply with any instructions. As the main difference between SP encounters and vignettes was that physicians knew that they were being observed when completing vignettes, comparing performance on vignettes with the performance of the same physician when treating SPs provides an estimate of the “know-do gap,” or the difference between what is done in practice relative to knowledge of correct procedures (Leonard and Masatu, 2005, 2010a; Mohanan et al., 2015). From these vignettes, we construct the same outcome measures as with SP visits.

### 3.3 Empirical Strategy

These data allow us to estimate two primary parameters of interest, which apply to two separate policy questions. One is the difference in the quality of care provided by physicians on fixed-term and civil service contracts, including differences in performance due to physician characteristics. This difference, referred to as the “as is” effect in the literature on paraprofessionals, addresses the question of how physicians in fixed-term contracts perform relative to civil service physicians *as they are currently hired*. A second question is the effect that the contract itself has on physician performance. This second parameter may be more relevant for contexts in which the interest is in whether physicians should be employed on fixed-term or civil service contracts. For this second parameter, the counterfactual is how physicians would perform if they had a fixed-term versus civil service contract.

The main difficulty in estimating both of these parameters is that fixed-term and civil-service physicians tend to be employed in different types of facilities and face different institutional structures and treat different types of patients as a result. The use of standardized patients allows us to address potential differences in patient and disease case mix because the cases presented to physicians are standardized and fixed by design (Das et al., 2016b). To address potential differences across facilities, we present estimates that control for facility fixed effects. Specifically, to estimate the “as is” difference between fixed-term and civil service physician performance, we estimate the following model, using ordinary least squares:

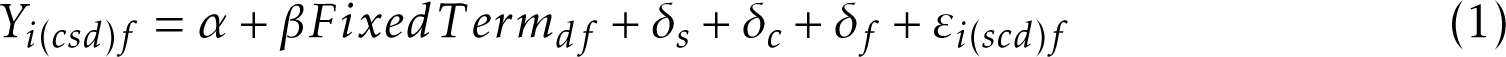

where *Y_i_*_(*scd*)*f*_, is the quality of care measure of interest in interaction *i* between SP *s* presenting case *c* and a physician *d* in facility *f* ; *FixedT erm_df_* is a dummy variable equal to one if the physician is on a fixed-term contract; *δ_s_* and *δ_c_* are SP and disease case fixed effects; and *ε_i_*_(*scd*)*f*_ a random error assumed to be unrelated to contract status. Of main interest is the coefficient *β*, which estimates the “as is” difference between fixed-term and civil service physicians. Our primary quality of care measure is the IRT-scaled score constructed from recommended checklist items. To aid interpretation, we normalize this score by the mean and standard deviation in the sample. In addition, we estimate differences in the probability of patients’ receiving a correct or partially correct treatment for the given disease case,^11^ whether patients are referred to upper-level facilities, total fees (including consultation and cost of drugs prescribed), the number of medicines prescribed, and whether any medicines were prescribed that are considered unnecessary or potentially harmful.

To estimate the effect of contract status, we re-estimate (1), including a vector of physician characteristics:

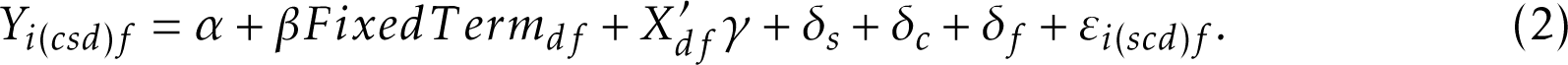

Here, *X_df_* includes physician age, sex, whether the physician has a medical degree from a vocational college or higher, whether the physician has a “practicing physician” certificate (the highest of three certifications), and rural household registration status (*hukou*). We also present estimates that additionally control for the physician’s IRT-scaled scores on clinical vignettes as well as indices of prosocial and intrinsic motivation. Clinical vignettes are matched to the some disease case presented to the same physician in the corresponding SP interaction. As these vignettes are designed to test competence, they should provide a reasonable proxy for disease-case-specific physician ability.

## 4 Results

### 4.1 The Quality of Care provided by Fixed-term and Civil Service Physicians

#### 4.1.1 Process Quality

The raw difference in performance between fixed-term and civil service physicians can be seen in Figure 1. Panel A shows a plot of the cumulative distribution functions (CDF) of the normalized IRT-scaled score based on clinical checklist completion in SP interactions for fixed-term and civil service physicians in the full sample. Panel B provides a plot of these functions for the sub-sample (38%) of facilities that employ both fixed-term and civil service physicians. In both cases, the distribution for fixed-term physicians stochastically dominates that for civil service physicians.^12^

**Figure 1:**
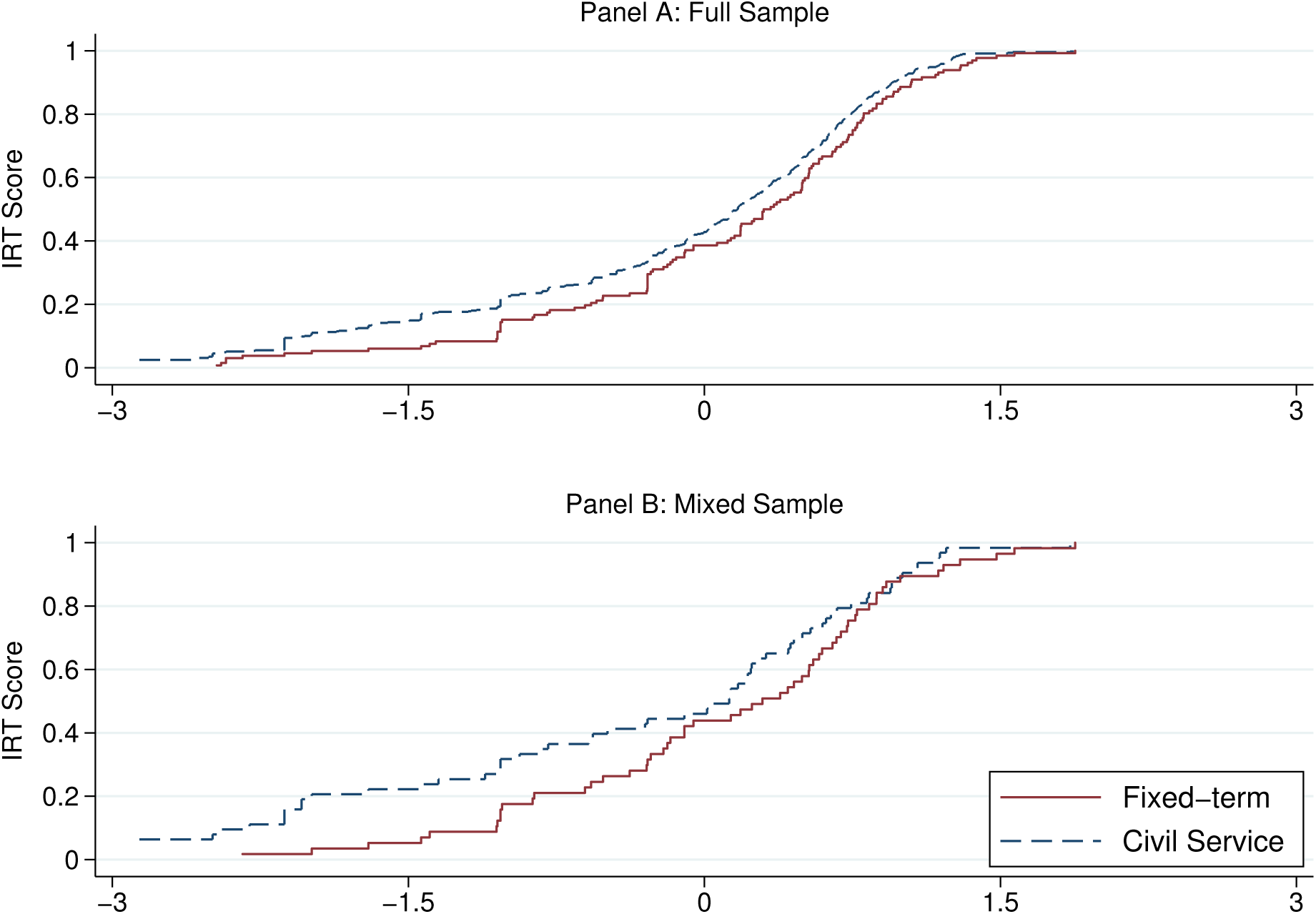
Cumulative Density Functions for Physician Process Quality

Table 3 presents the regressions that estimate this difference parametrically. The first three columns provide estimates for the “as is” effect of contract status. This is the difference in performance between contract and civil service physicians including the difference attributable to differences in observable characteristics. The second set of regressions control for observable physician characteristics, and therefore estimate the effect of the contract status alone.

**Table 3.**
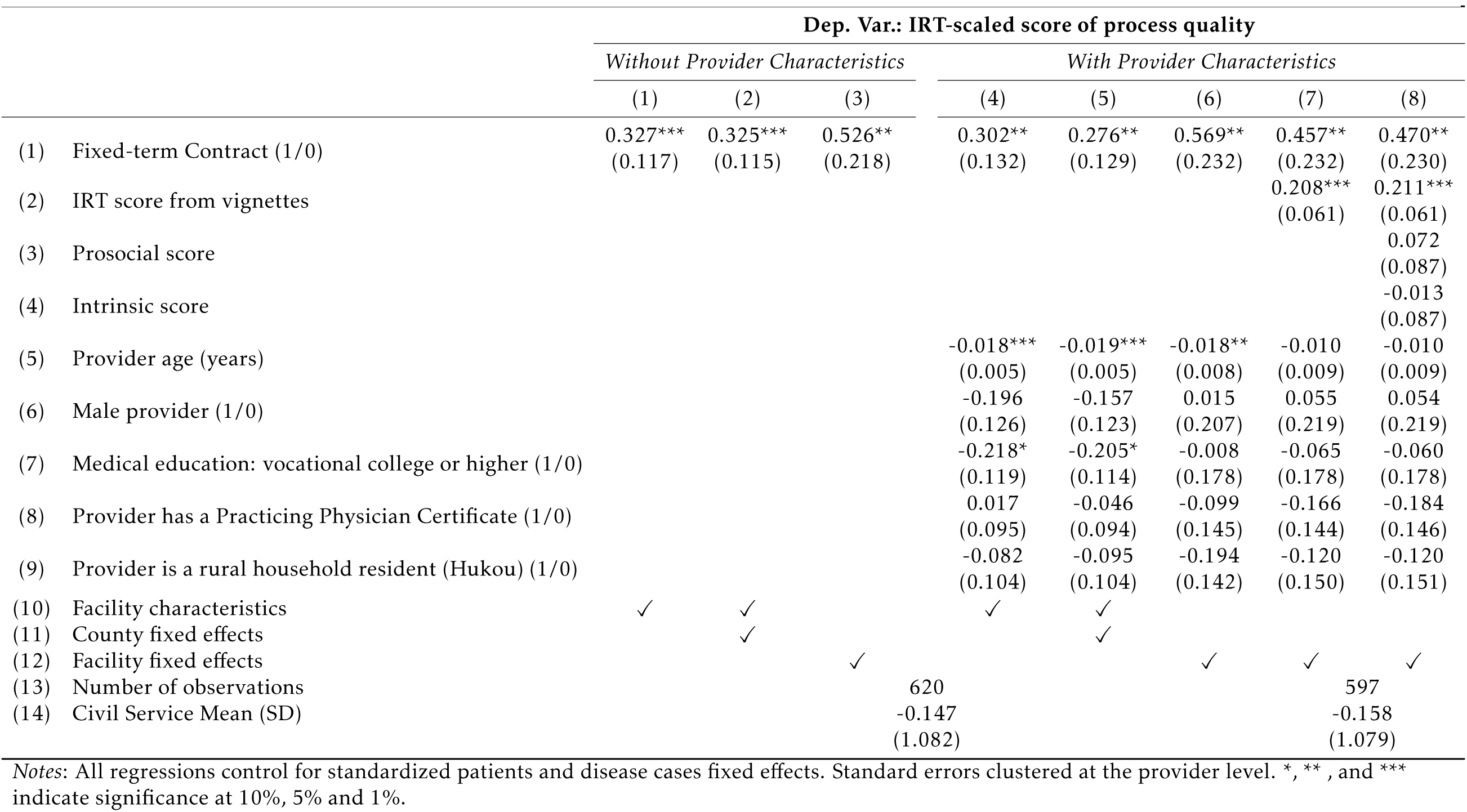
Physician Contract Status and Process Quality in SP Interactions

These results show that physicians on fixed-term contracts meaningfully outperform civil service physicians on process quality. Controlling only for observable facility characteristics and county fixed effects, we estimate that fixed-term physicians perform 0.33 standard deviations better on the IRT score compared to civil service physicians (Table 2, Column 2). In our specification that controls instead for facility fixed effects, the estimated “as is” difference increases to more than a one-half standard deviation (Column 3).

The second set of regressions, which control for observable physician characteristics, suggest that the nature of the contract itself has a large effect on process quality. Controlling for observable physician characteristics and facility fixed effects in the full sample, we estimate that the effect of the fixed-term relative to the civil service contract is 0.57 standard deviations (Column 6). Using the sample for which we have observations on the same physicians from both SP interactions and vignettes (597, 96% of the full sample), this estimate decreases slightly to 0.46 standard deviations when controlling for physician vignette scores (Column 7). Additionally controlling for prosocial and intrinsic motivation has little effect on the estimated impact of contract status.

#### 4.1.2 Treatment

In Table 4, we present estimates for our secondary outcomes related to treatment. We find no significant effects for either parameter on the probability of correct treatment, whether patients are referred to higher-level facilities, total fees charged, or drug prescriptions after controlling for facility fixed effects. There is suggestive evidence, however, that fixed-term contracts increase referrals and reduce unnecessary prescriptions and patient costs. We estimate meaningful effects on these three outcomes in specifications controlling for county fixed effects, finding an 11.5 percentage-point (46.6%) increase in referrals, 13.3 percentage-point (23.9%) reduction in unnecessary prescribing, and a 5.2 RMB (27.7%) reduction in costs (Column 5). These are no longer significant when controlling for facility fixed effects (Column 6), possibly due to reduced efficiency in these regressions, but we are able to reject that fixed-term contracts increase unnecessary drug prescriptions (one-sided *p*-value = 0.075). This finding is of note, as fixed-term positions are generally financed using revenue from the facility, and physicians employed on fixed-term contracts may face stronger incentives to provide unnecessary care. Nevertheless, more than half of both types of physicians in our sample prescribe medications that are unnecessary for a given disease case or are potentially harmful.

**Table 4.** Physician Contract Status and Treatment in SP interactions

|  | Without Provider Characteristics |  |  | With Provider Characteristics |  |  |
| --- | --- | --- | --- | --- | --- | --- |
|  | (1) | (2) | (3) | (4) | (5) | (6) |
| (1) Correct or partially treatment (1/0) | 0.015<br>(0.050) | 0.029<br>(0.050) | -0.073<br>(0.087) | 0.014<br>(0.056) | 0.023<br>(0.056) | -0.072<br>(0.104) |
| *Civil Service Mean* |  | 0.389 |  |  | 0.385 |  |
| (2) Referral (1/0) | 0.040<br>(0.050) | 0.065<br>(0.047) | -0.070<br>(0.069) | 0.094*<br>(0.051) | 0.115**<br>(0.049) | -0.047<br>(0.076) |
| *Civil Service Mean* |  | 0.258 |  |  | 0.247 |  |
| (3) Unnecessary/harmful medication (1/0) | -0.030<br>(0.052) | -0.046<br>(0.050) | -0.051<br>(0.086) | -0.110**<br>(0.054) | -0.133**<br>(0.053) | -0.142<br>(0.099) |
| *Civil Service Mean* |  | 0.541 |  |  | 0.555 |  |
| (4) Total cost (RMB) | -2.421<br>(2.260) | -2.863<br>(2.324) | -2.469<br>(3.829) | -4.825*<br>(2.838) | -5.210*<br>(2.878) | -3.353<br>(5.000) |
| *Civil Service Mean* |  | 18.327 |  |  | 18.778 |  |
| (5) Facility characteristics | ✓ | ✓ |  | ✓ | ✓ |  |
| (6) Provider characteristics |  |  |  | ✓ | ✓ | ✓ |
| (7) County fixed effects |  | ✓ |  |  | ✓ |  |
| (8) Facility fixed effects |  |  | ✓ |  |  | ✓ |
| (9) Number of observations |  | 620 |  |  | 597 |  |

Notes: All regressions control for standardized patients and disease cases fixed effects. \*, \*\*, and \*\*\* indicate significance at 10%, 5% and 1%.

### 4.2 Contract Status and the Know-do Gap

Vignettes that present the same disease case to the same physicians allow us to estimate the “know-do gap,” or difference between what is done in practice relative to knowledge of correct procedures. To compare the know-do gap between physicians employed on the two types of contracts, we stack observations from SP interactions and vignettes and estimate regression:

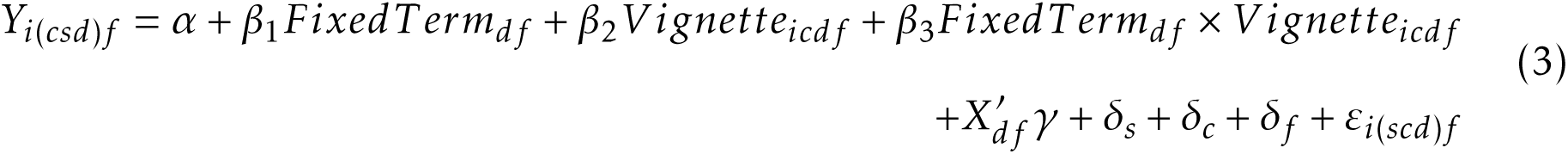

where *V ignette_icdf_* is a dummy variable for a vignette observation. *β*_2_ estimates the know-do gap among civil service physicians, *β*_2_ + *β*_3_ gives the know-do gap for fixed-term physicians, and *β*_3_ estimates the difference in know-do gaps between fixed-term and civil service physicians. We estimate this with and without facility fixed effects and with and without the full set of physician characteristics for all outcomes, but exclude total fees, as fees are not available for vignette observations. We also replace the IRT-scaled process quality score with the percentage of checklist items completed, as this measure is absolute (rather than relative) and directly comparable across SP interactions and vignettes.

The know-do gap is significantly smaller for fixed-term physicians on all dimensions, except unnecessary medications, with and without controlling for observable characteristics (Table 5). In terms of process quality, we find a significant know-do gap for civil service physicians but not for fixed-term physicians (Panel A). ^13^ On average, the same civil service physicians complete around 4.6% fewer recommended checklist items in interactions with unannounced SPs compared to when presented with identical disease cases as vignettes. No such gap is present for fixed-term physicians. Further, although both types of physicians are more likely to provide correct or partially correct treatments in vignettes than in SP interactions, this gap is a full 10 percentage points larger for civil service physicians (Panel B). Civil servants also are significantly more likely to suggest referral to higher-level hospitals in vignette cases compared to when presented with unannounced SPs (Panel C). We do not find any evidence of know-do gaps for either type of physician in terms of prescription practices.

**Table 5.**
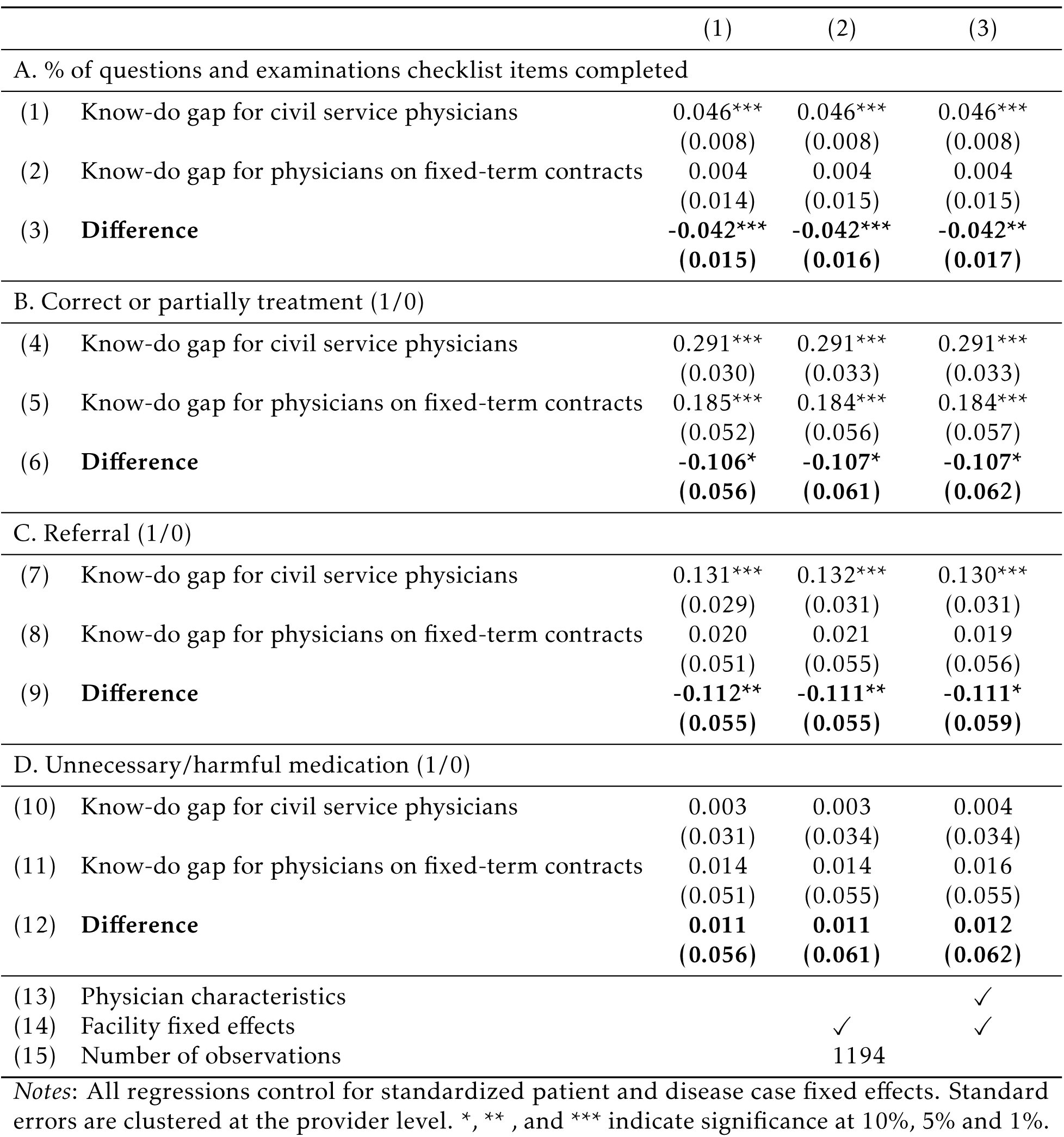
Physician Contract Status and the Know-do Gap

In sum, the estimates show that, although civil service physicians tend to have slightly higher levels of knowledge of appropriate clinical practice, fixed-term physicians perform closer to their ability/knowledge (i.e., have smaller know-do gaps). This implies that the differences that we find in the performance of the two types of physicians are almost entirely driven by differences in effort.

### 4.3 Heterogeneous Effects: Ability and Internal Motivation

Fixed term contracts appear to elicit greater effort than civil service contracts among physicians in our data, but effects may depend on physician characteristics. If so, how contracts effect performance could interact with policies that influence the composition of the public-sector physician workforce. Such policies include both those affecting the skills and motivation of physicians already employed (such as training) and how physicians select into public service. In addition to selection effects of job attributes, recruitment policies intentionally or consequently target physicians with specific characteristics, such as high levels of prosocial or mission-oriented motivation (Besley and Ghatak, 2005; Ashraf et al., 2019).

We estimate how physician ability, prosocial motivation, and intrinsic motivation interact with contract status by estimating:

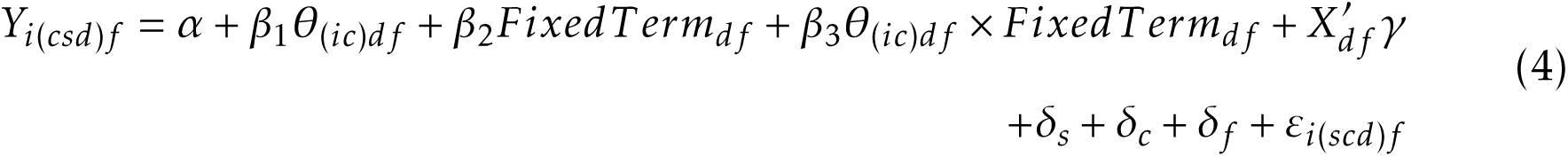

where *θ_df_* is the normalized process score from clinical vignettes, an index for prosocial motivation, or an index for intrinsic motivation. We allow for non-linear effects in skills and motivation by defining *θ* as dummy variables that indicate values above the median in the sample.^14^ All regressions control for the full set of physician characteristics and facility, SP, and disease-case fixed effects.

In Table 6, we present three quantities for each outcome: effect of contract status on physicians with high levels of the characteristic (*β*_2_ + *β*_3_, first row in each panel), low levels of the characteristic (*β*_2_, second row), and the p-value corresponding to the estimate for *β*_3_ (third row). We do not find strong evidence that the effect of contract status varies with physician ability/skills for any of the outcomes. The estimated effect of fixed-term contracts on process quality (as measured in SP interactions) among low-ability physicians is larger than that for high-ability physicians (0.72 vs. 0.32 SD), but the difference in effects is not significant.

**Table 6.** Heterogeneous Effects of Contract Status on Diagnostic Process Quality in SP Interactions

|  |  | IRT-scaled score of<br>process quality | Correct or partially<br>treatment (1/0) | Referral (1/0) | Unnecessary/harmful<br>medication (1/0) | Total cost |
| --- | --- | --- | --- | --- | --- | --- |
| A. Ability (high = vignette IRT score > median) |  | (1) | (2) | (3) | (4) | (5) |
| (1) Fixed-term effect among high ability |  | 0.317<br>(0.299) | -0.138<br>(0.138) | -0.014<br>(0.106) | -0.182<br>(0.136) | -3.654<br>(5.561) |
| (2) Fixed-term effect among low ability |  | 0.722***<br>(0.265) | -0.017<br>(0.117) | -0.072<br>(0.086) | -0.122<br>(0.110) | -4.373<br>(6.241) |
| (3) *P-value: Interaction Term* |  | 0.196 | 0.416 | 0.605 | 0.682 | 0.916 |
| B. Prosocial motivation (high = index > median) |  | (1) | (2) | (3) | (4) | (5) |
| (4) Fixed-term effect among high prosocial |  | 0.662**<br>(0.270) | -0.145<br>(0.124) | -0.025<br>(0.099) | -0.031<br>(0.115) | 0.882<br>(6.513) |
| (5) Fixed-term effect among low prosocial |  | 0.044<br>(0.305) | 0.086<br>(0.167) | -0.027<br>(0.101) | -0.174<br>(0.127) | -5.123<br>(5.514) |
| (6) *P-value: Interaction Term* |  | 0.074 | 0.231 | 0.991 | 0.351 | 0.418 |
| C. Intrinsic motivation (high = index > median) |  | (1) | (2) | (3) | (4) | (5) |
| (7) Fixed-term effect among high intrinsic |  | 0.558**<br>(0.290) | -0.150<br>(0.117) | -0.136<br>(0.095) | -0.103<br>(0.128) | -0.303<br>(6.486) |
| (8) Fixed-term effect among low intrinsic |  | 0.313<br>(0.259) | 0.057<br>(0.164) | 0.113<br>(0.115) | -0.217*<br>(0.129) | -8.786<br>(5.423) |
| (9) *P-value: Interaction Term* |  | 0.451 | 0.244 | 0.080 | 0.497 | 0.231 |
| (10) Number of observations |  | 597 |  |  |  |  |

Notes: All regressions control for the full set of provider characteristics and facility, standardized patient and disease case fixed effects. Standard errors, in parentheses, are clustered at the provider level. \*, \*\*, and \*\*\* indicate significance at 10%, 5% and 1%.

Effects of contract status do, however, vary with social preferences. First, we find evidence that fixed-term contracts may increase effort by crowding-in prosocial motivation. The results in Column 1 of Table 7 indicate that the effect of fixed-term contracts on process quality is 0.62 standard deviations larger among physicians with higher levels of prosocial motivation. Assuming linearity, we estimate that the effect of the fixed-term contract increases by 0.21 standard deviations with a 1 standard deviation increase in the prosocial index (Appendix Table 5, Column 1).

**Table 7.** Correlates of Physician Pay

|  |  | Dep. Var.: ln(Physician Income) |  |  |
| --- | --- | --- | --- | --- |
|  |  | Full Sample | Civil Service Physicians | Fixed-term Physicians |
| A. Physician Characteristics |  | (1) | (2) | (3) |
| (1) | Experience at this facility (years) | 0.009**<br>(0.004) | 0.007*<br>(0.004) | 0.027*<br>(0.015) |
| (2) | Experience at this facility (years)<sup>2</sup> | -0.000<br>(0.000) | -0.000<br>(0.000) | -0.001**<br>(0.000) |
| (3) | Education, upper secondary or higher (1/0) | 0.041<br>(0.028) | 0.007<br>(0.030) | 0.016<br>(0.099) |
| (4) | Provider has a Practicing Physician Certificate (1/0) | 0.178***<br>(0.031) | 0.099***<br>(0.033) | 0.276**<br>(0.122) |
| (5) | Male (1/0) | 0.042<br>(0.035) | 0.035<br>(0.038) | 0.058<br>(0.110) |
| (6) | Provider is a rural household resident (Hukou) (1/0) | -0.086***<br>(0.031) | -0.040<br>(0.031) | 0.044<br>(0.099) |
| (7) | Average IRT Score from vignettes | 0.009<br>(0.014) | 0.022<br>(0.015) | 0.043<br>(0.034) |
| (8) | Prosocial score | 0.009<br>(0.016) | 0.022<br>(0.017) | -0.055<br>(0.054) |
| (9) | Intrinsic score | -0.011<br>(0.017) | -0.018<br>(0.017) | 0.041<br>(0.079) |
| B. Facility Characteristics |  |  |  |  |
| (10) | Number of patients in catchment area (1000 patients) | -0.000<br>(0.001) | -0.000<br>(0.001) | -0.002<br>(0.004) |
| (11) | Number of civil service doctors at the facility | -0.001<br>(0.003) | -0.001<br>(0.003) | -0.005<br>(0.011) |
| (12) | Total values of equipment (10,000 Yuan) | -0.000<br>(0.000) | 0.000<br>(0.000) | -0.001<br>(0.002) |
| (13) | Central Township Health Center (1/0) | 0.035<br>(0.035) | 0.021<br>(0.035) | 0.134<br>(0.095) |
| (14) | Number of village clinics in this township | 0.002<br>(0.002) | 0.002<br>(0.002) | 0.003<br>(0.004) |
| (15) | Distance to nearest county hospital (kilometers) | 0.000<br>(0.001) | -0.001<br>(0.001) | 0.002<br>(0.002) |
| (16) | Net Revenue (10,000 Yuan) | 0.000<br>(0.000) | 0.000<br>(0.000) | 0.000<br>(0.001) |
| (17) | $R^2$ | 0.52 | 0.64 | 0.58 |
| (18) | Number of observations | 370 | 293 | 77 |

Notes: All regressions control for county fixed effects. Standard errors are clustered at the facility level. \*, \*\*, and \*\*\* indicate significance at 10%, 5% and 1%.

We also find suggestive evidence that the relationship between prosocial motivation and prescription of unnecessary medications to SPs varies with contract status. Among civil service physicians, having a high prosocial score is associated with a decrease in unnecessary prescriptions by 31 percentage points, representing a substantial 54% decrease. The estimated relationship among fixed-term physicians, however, is 14 percentage points smaller and insignificant. Although the difference between effects is insignificant in the non-linear specifications (Table 7, Panel B, Column 4), the interaction between contract status and the prosocial index is significant assuming a linear relationship (entering the index score as a continuous variable; Appendix Table 5, Column 4).

A question that arises from these results is why extrinsic incentives that stem from fixed-term contracts do not appear to crowd-out prosocial motivation. Although untestable with our data, one hypothesis is that this has to do with eroded professional norms among physicians in China. Bénabou and Tirole (2006) present a model in which crowding-out is likely to occur when extrinsic incentives create doubt about the true motive for good deeds. In other words, extrinsic rewards may crowd-out prosocial motivation when they “spoil” the image that effort is due to the altruism of an individual. In our context, the potential for crowding-out would thereby depend on the degree to which physicians value a prosocial image. Even when physicians are prosocially motivated, extrinsic rewards may not crowd-out effort if a prosocial image is not valued within an organization. In China, it has been argued that professional norms and ethics among physicians have been eroded due to the abolishment of professional organizations and decades of perverse incentives tied to explicit profit motives (Yip et al., 2010). If the value of having a prosocial image is similarly degraded, stronger extrinsic incentives may not have the same potential to crowd-out internal motivation as they would compared to settings with stronger professional norms.

### 4.4 Determinants of Civil Service and Fixed-term Pay

Civil service physicians may face weak incentives relative to fixed-term physicians for at least two reasons. One explanation is that, as dismissal is rare and promotions near automatic, career concerns do not provide strong incentives in civil service. Once hired, promotions are based primarily on qualifications and experience. For fixed-term employees, career concerns may be stronger, due not only to the possibility of dismissal but also to the potential for eventual civil service employment (and better non-pecuniary benefits) (Duflo et al., 2015).

A second explanation is that pay does not reward better performance in civil service but does in the market for fixed-term physicians. It has been argued that the civil service headcount quota (*bianzhi*) system creates misallocation in the labor market for physicians by limiting mobility (World Bank, 2019). Fixed-term contracts may therefore create stronger incentives by allowing the market to effectively link pay to performance. Of course, this depends on whether better quality of care, or something correlated with this dimension of performance, is valued by managers and is observable. In India, Das et al. 2016b find that pricing of primary care in the private sector is related to performance, suggesting that quality is, at least in part, observable to consumers.

To explore these two mechanisms, we examine the relationships between physician characteristics and pay, and between pay and observed quality (as measured using outcomes from SP interactions) differ by physician type.

#### 4.4.1 Physician Characteristics and Earnings

We estimate three Mincer equations relating physician characteristics and pay for the full sample (combining both physician types), civil service physicians only, and fixed-term physicians only:

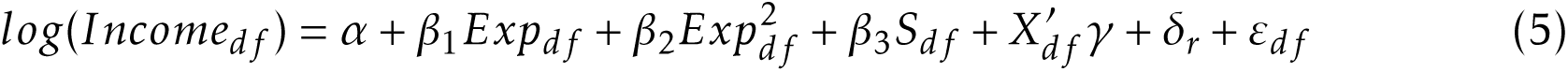

where *Income_df_* is the monthly income of physician *d* in facility *f* ; *Exp_df_* is years working in the same facility; *S_df_* is whether the physician has a secondary education or higher; *X_df_* is a vector of additional physician (certification, gender, rural resident status (*hukou*), and average IRT score in vignettes as an ability proxy) and facility characteristics (same as Appendix Table 1); and *δ_r_* are county fixed effects. Standard errors account for clustering within facilities.

There are two findings of note from these regressions (Table 7). First, experience and qualifications appear to be more strongly rewarded in the market for fixed-term physicians. The relationship between experience and earnings among fixed-term employees is more than three times larger than for civil service employees. The premium for fixed-term physicians with a practicing physician certificate is also nearly three times larger than the premium for civil service physicians. Second, differences in the *R*^2^s in each regression show that experience and qualifications explain a larger portion of the variation in pay across civil service physicians (0.64 vs. 0.58).

Taken together, these regressions suggest that civil servant pay is somewhat more deterministic and depends primarily on tenure and qualifications. Though fixed-term physicians are more strongly rewarded for experience and qualifications, there is also more room for other factors, such as demonstrated effort, to affect earnings.

#### 4.4.2 Pay and Performance

To test this link between pay and performance, we explore the relationship between performance scores from interactions with SPs and physician pay for both physician types. Figure 2 shows the non-parametric relationship between IRT-scaled scores in interactions with SPs and physician income along with the 90% confidence intervals. We see that civil service income is uncorrelated with the performance score. In contrast, fixed-term physician income appears to have a positive relationship with quality.

**Figure 2:**
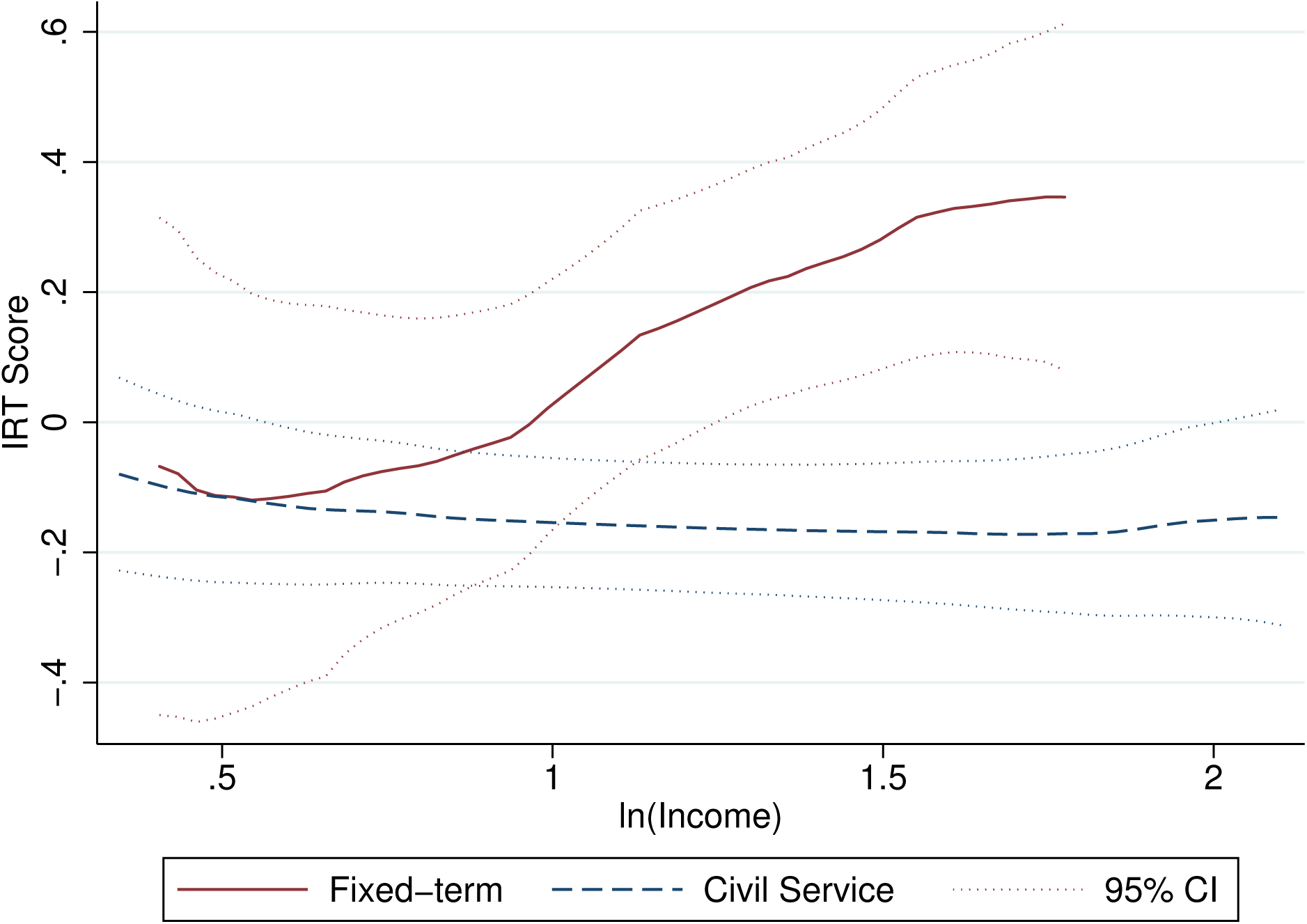
Physician Pay and Performance

We estimate this relationship parametrically using variants of the following specification:

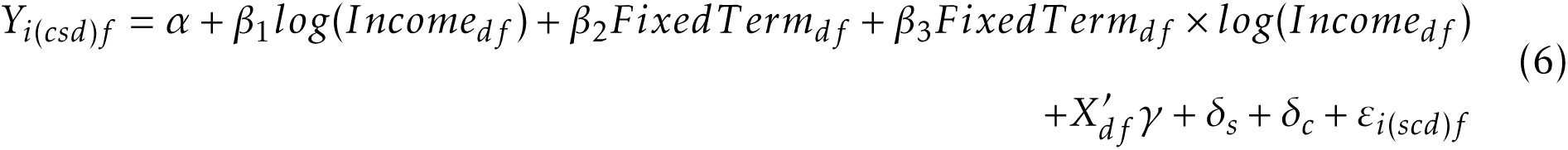

where *Y_i_*_(*scd*)*f*_ is the normalized IRT-scaled score from interactions with SPs; *X_df_* progressively includes physician and facility characteristics as well as city or county-level fixed effects; and all else is defined as in previous regressions. Here, *β*_1_ is the estimated correlation between pay and performance for civil service physicians and *β*_1_ + *β*_3_ gives the correlation among fixed-term physicians.

The results in Table 8 show that the same pattern holds as we control for other observable characteristics: In all specifications, civil service pay is unrelated to the performance measure, whereas there is a significant correlation among fixed-term physicians in the first four specifications. The difference in the two correlations is significant in all but the final specification, controlling for county fixed effects. That the correlation between pay and income for fixed-term physicians shrinks when controlling for city and county fixed effects (Columns 4 and 5) suggests that this relationship may be driven, in part, by mobility between cities and counties although we cannot rule out other factors.

**Table 8.** The Correlation Between Physician IRT Score, Income and Contract Status

|  |  | Dep. Var.: IRT-scaled score of process quality |  |  |  |  |
| --- | --- | --- | --- | --- | --- | --- |
|  |  | (1) | (2) | (3) | (4) | (5) |
| (1) | ln(Physician Income) | -0.089<br>(0.164) | -0.118<br>(0.159) | -0.107<br>(0.168) | -0.017<br>(0.200) | -0.034<br>(0.201) |
| (2) | Fixed-term | -0.446<br>(0.372) | -0.486<br>(0.389) | -0.453<br>(0.379) | -0.307<br>(0.419) | -0.217<br>(0.419) |
| (3) | ln(Physician Income) * Fixed-term | 0.632**<br>(0.302) | 0.670**<br>(0.317) | 0.639**<br>(0.304) | 0.520<br>(0.329) | 0.466<br>(0.322) |
| (4) | **Correlation among fixed-term physicians (Row 1 + Row 3)** | **0.543\*\***<br>**(0.245)** | **0.552\*\***<br>**(0.268)** | **0.532\*\***<br>**(0.264)** | **0.503\***<br>**(0.265)** | **0.433**<br>**(0.265)** |
| (5) | Physician characteristics |  | ✓ | ✓ | ✓ | ✓ |
| (6) | Facility characteristics |  |  | ✓ | ✓ | ✓ |
| (7) | City fixed effects |  |  |  | ✓ |  |
| (8) | County fixed effects |  |  |  |  | ✓ |
| (9) | Number of observations | 620 |  |  | 597 |  |

Notes: All regressions control for disease case fixed effects. Standard errors are clustered at the facility level. \*, \*\*, and \*\*\* indicate significance at 10%, 5% and 1%.

While correlations, these estimates provide suggestive evidence that, in addition to stronger or more salient career concerns, fixed-term physicians’ incentives may be better aligned with quality because their pay is linked to performance. That civil servant pay is not related to performance is in line with the previous findings for public sector physicians in India (Das et al., 2016b) and teachers in Pakistan (Bau and Das, 2017), but in our context it seems that physicians are rewarded for better performance when employed outside of civil service posts even though still in the public sector.

## 5 Conclusion

Over the past several decades, multiple countries have sought to improve the delivery of publicly provided services by reforming civil service employment. Driven by the hypothesis that rigid rules that govern public-sector employment weaken the incentives of those who provide services, a prominent feature of these reforms has been to grant greater decision-making power to facility managers over hiring and firing and employee compensation. We contribute evidence relevant to these reforms by exploring the performance of civil service physicians and physicians employed on fixed-term contracts in China. The institutional setting combined with our data collection, using audits with matching clinical vignettes, allows us to address important sources of endogeneity in estimating the effects of contract status.

We present five main findings. First, physicians employed on fixed-term contracts substantially outperform physicians employed as civil servants on measures of process quality. This is the case even though physicians employed on fixed-term contracts tend to have lower levels of education and certification.

Second, despite the potential for stronger incentives to generate clinic revenue, we find no evidence that physicians on fixed-term contracts are more likely to provide unnecessary treatments. This is of note as fixed-term positions are financed through clinic revenues and given concerns in China that physicians generally have faced incentives for unnecessary care and waste as a result of clinic revenues tied to drug sales (Yip et al., 2010).

Third, our analysis suggests performance differences arise due to effects of contract status on effort. Although civil service physicians exhibit slightly better knowledge of appropriate clinical practice than do fixed-term physicians, they perform substantially worse relative to this knowledge.

Fourth, we find that the effects of contract status vary with prosocial motivation. The effects of contract status on effort are stronger among those with higher levels of prosocial motivation, as measured using psychological scales. We also find some evidence that contract status may mediate the relationship between prosocial motivation and unnecessary treatment. The interaction between prosocial motivation and contract status is important in light of policies that affect how physicians select into public service. In China, ongoing reforms to the medical education system and initiatives to recruit medical students from rural areas, for example, likely affect the composition of the physician workforce on a number of dimensions, including levels of internal motivation.

Finally, our analysis of physician pay shows that, although pay is unrelated to performance for civil service, there is a significant relationship for fixed-term physicians. Fixed-term contracts may, therefore, strengthen incentives both through career concerns and because fixed-term physicians are effectively rewarded for better performance with higher pay.

There are several limitations to this study. Although we are able to address some important sources of endogeneity, several threats to internal and external validity remain. First, even after controlling for a large set of observable characteristics (including a measure of physician ability), unobserved differences may remain that bias estimates away from the true effect of contract status. Although we cannot rule this out completely, calculations following Oster (2016), suggest that unoberservables would need to be 1.47 times as important as observables included in the main specification for the true effect of contract status on process quality to be zero.^15^

There are four main threats to external validity. First, the SP and vignette disease case scripts present three diseases (diarrhea, tuberculosis, and unstable angina), which, while chosen to span important disease categories, may not be representative of diseases seen in outpatient settings more generally. Second, we drew our sample from rural areas in three provinces in Eastern, Central, and Western China to increase geographic diversity of the sample; however, these provinces were not randomly sampled and cannot be said to represent the entire country. Third, as with most quasi-experimental studies, our estimates are “local” in that they are identified off variation in a subsample of the data, in our case the 38% of facilities that employed both types of physicians. As a result, we cannot say whether results would be the same in facilities that do not currently employ fixed-term physicians.

An additional threat corresponds to the plausibility of the Stable Unit Treatment Value Assumption (SUTVA) (Cox, 1958; Rubin, 1980). There are two potential concerns: First, we estimate the effect of fixed-term vs. civil service contracts including other attributes of these employment contracts (e.g., pay, bonus structure, fringe benefits, opportunities for career advancement, etc). Our analysis may therefore not correspond, for example, to shifting civil service physicians to fixed term contracts if these other attributes also change. Second, we are unable to rule out the possibility of peer effects between physicians in the same facility. Assuming no such spillovers exist, our estimates would correspond to both the current setting where both civil service and fixed-term contracts are used as well as to a policy abolishing civil service positions for physicians entirely. Two possibilities for such spillovers include (1) direct peer effects (e.g. more effort among fixedterm physicians increases the effort of civil service physicians) and (2) because fixed-term physicians are financed with facility revenue, civil service physicians may face stronger incentives to generate revenue through drug sales or other means. To explore this possibility, Appendix Table 6 shows estimates for the relationship between whether a facility hires any fixed-term physicians and the outcomes from standardized patient visits. We do not find any evidence that hiring fixed term physicians is related to the process quality of civil service physicians; however there is some evidence that civil service physicians may respond by generating more clinic revenue through prescriptions of unnecessary drugs and less frequently referring patients to higher-level facilities. When instrumenting for hiring of fixed-term physicians with the ratio of population in the catchment area to civil service posts allocated the clinic, we estimate that civil service physicians reduce referrals by 25 percentage points. Similar estimates for patient out-of-pocket costs and unnecessary drug prescription are positive and large, but insignificant.

Despite these limitations, our results provide strong evidence that a fundamental issue that affects the quality of services provided by public facilities may be civil service regulations that limit manager autonomy over personnel decisions. Important areas for future study include experimental work to identify the causal effect of civil service employment relative to alternative forms of physician employment in China and to compare these with other contract features, such as the use of performance pay.

## Data Availability

Data and code for replication available in the UNC Dataverse

https://doi.org/10.15139/S3/YGO7DT

---

**Appendix Table 1.**
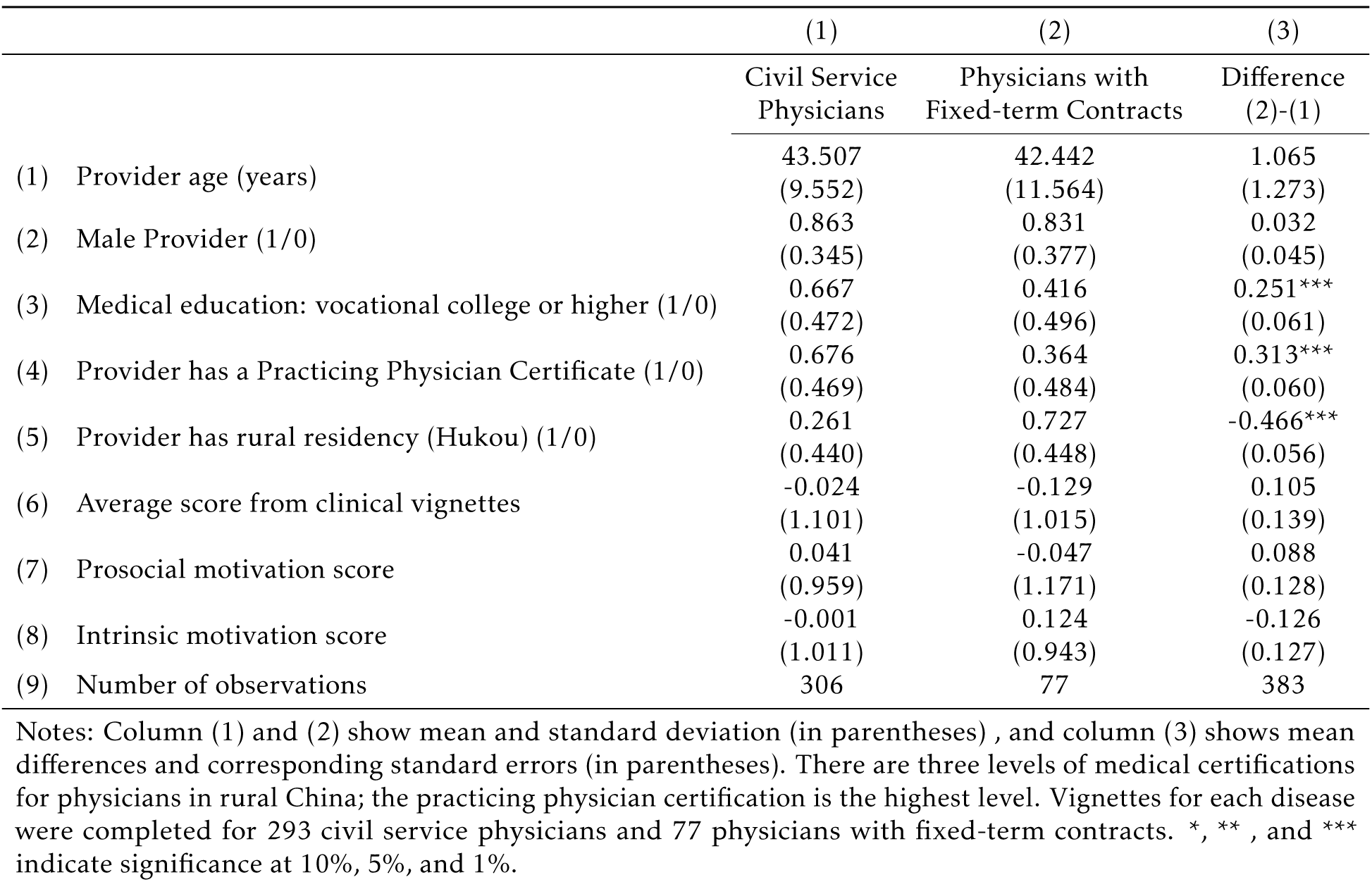
Physician Characteristics (Performance Sample)

**Appendix Table 2.**
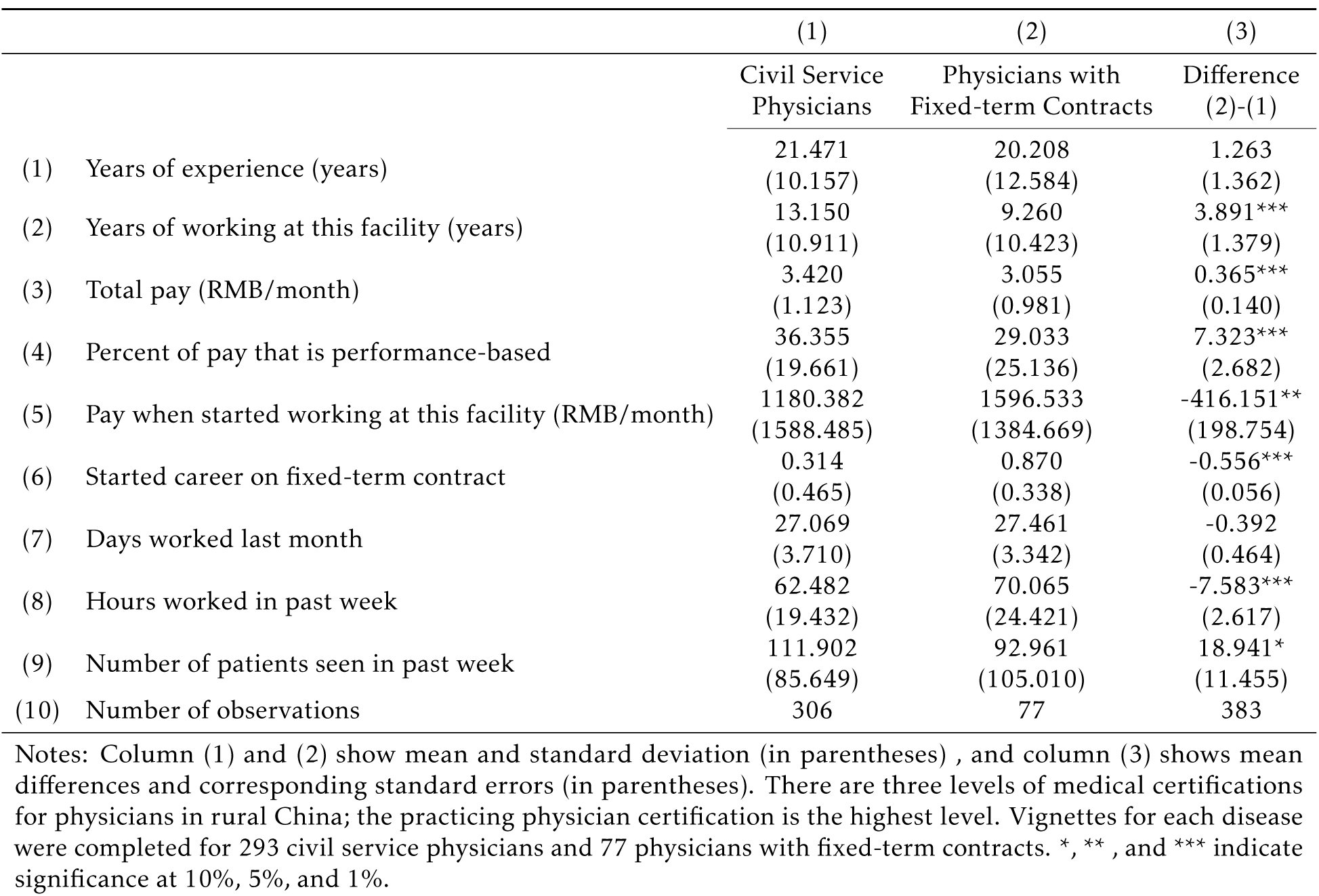
Physician Experience, Pay, and Workload (Performance Sample)

|  | (1) | (2) | (3) |
| --- | --- | --- | --- |
|  | Civil Service<br>Physicians | Physicians with<br>Fixed-term Contracts | Difference<br>(2)-(1) |
| (1) Years of experience (years) | 21.471<br>(10.157) | 20.208<br>(12.584) | 1.263<br>(1.362) |
| (2) Years of working at this facility (years) | 13.150<br>(10.911) | 9.260<br>(10.423) | 3.891***<br>(1.379) |
| (3) Total pay (RMB/month) | 3.420<br>(1.123) | 3.055<br>(0.981) | 0.365***<br>(0.140) |
| (4) Percent of pay that is performance-based | 36.355<br>(19.661) | 29.033<br>(25.136) | 7.323***<br>(2.682) |
| (5) Pay when started working at this facility (RMB/month) | 1180.382<br>(1588.485) | 1596.533<br>(1384.669) | -416.151**<br>(198.754) |
| (6) Started career on fixed-term contract | 0.314<br>(0.465) | 0.870<br>(0.338) | -0.556***<br>(0.056) |
| (7) Days worked last month | 27.069<br>(3.710) | 27.461<br>(3.342) | -0.392<br>(0.464) |
| (8) Hours worked in past week | 62.482<br>(19.432) | 70.065<br>(24.421) | -7.583***<br>(2.617) |
| (9) Number of patients seen in past week | 111.902<br>(85.649) | 92.961<br>(105.010) | 18.941*<br>(11.455) |
| (10) Number of observations | 306 | 77 | 383 |

Notes: Column (1) and (2) show mean and standard deviation (in parentheses) , and column (3) shows mean differences and corresponding standard errors (in parentheses). There are three levels of medical certifications for physicians in rural China; the practicing physician certification is the highest level. Vignettes for each disease were completed for 293 civil service physicians and 77 physicians with fixed-term contracts. \*, \*\*, and \*\*\* indicate significance at 10%, 5%, and 1%.

**Appendix Table 3.** Correlation between Facility Characteristics and Employment of Fixed-term Physicians

|  | Facilities with-<br>out Fixed-term<br>Physicians | Facilities with<br>Fixed-term<br>Physicians | Difference<br>(2)-(1) | Dep. Var.:<br>Facility Employs<br>Fixed-term Physi-<br>cians |
| --- | --- | --- | --- | --- |
|  | (1) | (2) | (3) | (4) |
| (1) Number of patients in catchment area (1000 patients) | 29.533<br>(22.230) | 20.898<br>(15.680) | 8.635***<br>(2.847) | -0.008***<br>(0.002) |
| (2) Number of civil service positions at facility | 6.914<br>(5.804) | 5.537<br>(6.675) | 1.377<br>(0.877) | -0.006<br>(0.006) |
| (3) Total value of equipment (10,000 Yuan) | 65.073<br>(66.820) | 66.456<br>(61.486) | -1.383<br>(9.239) | 0.001*<br>(0.001) |
| (4) Central Township Health Center (1/0) | 0.461<br>(0.500) | 0.525<br>(0.503) | -0.064<br>(0.071) | 0.115<br>(0.072) |
| (5) Number of village clinics in township | 14.633<br>(11.286) | 16.387<br>(12.103) | -1.755<br>(1.654) | 0.006**<br>(0.003) |
| (6) Distance to nearest county hospital (kilometers) | 29.820<br>(16.402) | 30.600<br>(18.131) | -0.780<br>(2.435) | -0.001<br>(0.002) |
| (7) Net Revenue (10,000 Yuan) | 7.222<br>(70.491) | 11.341<br>(76.222) | -4.119<br>(10.367) | 0.000<br>(0.000) |
| (8) Number of observations | 128 | 80 | 208 |  |
| (9) Adjusted R<sup>2</sup> |  |  |  | 0.07 |

Notes: Column (1) and (2) show mean and standard deviations (in parentheses) and column (3) shows mean differences and corresponding standard errors. Column (4) shows coefficients from an OLS regression predicting employment of fixed-term physicians at the facility. \*, \*\*, and \*\*\* indicate significance at 10%, 5%, and 1%.

**Appendix Table 4.** Standards for Correct, Partially Correct and Incorrect Treatments

|  | **Diarrhea** | **Unstable Angina** | **Tuberculosis** |
| --- | --- | --- | --- |
| Correct | ORS (Home-made ORS),<br>+/-Zinc,<br>No antibiotics,<br>No anti-diarrhea drugs | At least one of the six types of drugs;<sup>a</sup><br>Referral,<br>No antibiotics | A chest radiograph [CXR], or spu-<br>tum smear test, or referral,<br>No antibiotics.<br>No steroid |
| Partially Correct | ORS (Home-made ORS),<br>+/-Zinc, | At least one of the six types of drugs;<sup>a</sup><br>Referral | A chest radiograph [CXR], or spu-<br>tum smear test, or referral, |

<sup>a</sup> Six types of drugs for unstable angina: aspirin, anti-platelet, statins, nitroglycerin, beta-blockers, and ACEI (Angiotensin Converting Enzyme Inhibitors).

**Appendix Table 5.** Heterogeneous Effects of Contract Status on Diagnostic Process Quality in SP Interactions (Linear)

|  | *IRT-scaled score of*<br>*process quality*<br>(1) | *Correct or partially*<br>*treatment (1/0)*<br>(2) | *Referral (1/0)*<br>(3) | *Unnecessary/harmful*<br>*medication (1/0)*<br>(4) | *Total cost*<br>(5) |
| --- | --- | --- | --- | --- | --- |
| (1) IRT score from vignette survey | 0.237***<br>(0.070) | 0.024<br>(0.038) | -0.002<br>(0.026) | 0.012<br>(0.034) | -0.977<br>(1.637) |
| (2) Fixed-term | 0.446*<br>(0.235) | -0.076<br>(0.105) | -0.050<br>(0.078) | -0.157<br>(0.099) | -3.955<br>(4.757) |
| (3) IRT score from vignette survey * Fixed-term | -0.124<br>(0.147) | -0.023<br>(0.074) | -0.016<br>(0.052) | -0.077<br>(0.065) | -3.155<br>(2.780) |
| (4) Prosocial score | 0.002<br>(0.096) | -0.029<br>(0.051) | -0.054<br>(0.037) | -0.135***<br>(0.049) | -5.102*<br>(2.966) |
| (5) Fixed-term | 0.465**<br>(0.229) | -0.070<br>(0.106) | -0.047<br>(0.076) | -0.146<br>(0.098) | -3.402<br>(4.965) |
| (6) Prosocial score * Fixed-term | 0.210*<br>(0.123) | -0.080<br>(0.079) | -0.002<br>(0.048) | 0.126**<br>(0.058) | 1.862<br>(3.421) |
| (7) Intrinsic score | -0.035<br>(0.090) | 0.061<br>(0.054) | 0.044<br>(0.037) | 0.088*<br>(0.050) | 7.787***<br>(2.810) |
| (8) Fixed-term | 0.439**<br>(0.223) | -0.058<br>(0.108) | -0.032<br>(0.078) | -0.161*<br>(0.097) | -3.830<br>(4.932) |
| (9) Intrinsic score * Fixed-term | 0.156<br>(0.134) | -0.070<br>(0.079) | -0.078<br>(0.073) | 0.094<br>(0.071) | 2.416<br>(4.202) |
| (10) Number of observations | 597 |  |  |  |  |

*Notes:* All regressions control for provider characteristics, facility fixed effects, standardized patient and disease case fixed effects. Standard errors are clustered at the provider level. \*, \*\*, and \*\*\* indicate significance at 10%, 5% and 1%.

**Appendix Table 6.** Fixed-term Physician Hiring and Spillovers

|  | Full Sample (Civil Service & Fixed-term)<br>(N=204) |  |  | Civil Service Only<br>(N=183) |  |  |
| --- | --- | --- | --- | --- | --- | --- |
|  | OLS | RF | IV | OLS | RF | IV |
|  | (1) | (2) | (3) | (4) | (5) | (6) |
| *A. Process Quality Score (IRT-scaled)* |  |  |  |  |  |  |
| (1) Facility hired any fixed-term physicians | -0.072<br>(0.110) |  | 0.170<br>(0.250) | -0.181<br>(0.128) |  | -0.001<br>(0.467) |
| (2) Population per civil service post (1,000) |  | 0.006<br>(0.010) |  |  | -0.000<br>(0.024) |  |
| *B. Cost* |  |  |  |  |  |  |
| (3) Facility hired any fixed-term physicians | 3.286<br>(2.181) |  | -6.930<br>(7.659) | 5.505**<br>(2.477) |  | 7.279<br>(8.744) |
| (4) Population per civil service post (1,000) |  | -0.269<br>(0.279) |  |  | 0.351<br>(0.455) |  |
| *C. Unnecessary/harmful medication (1/0)* |  |  |  |  |  |  |
| (5) Facility hired any fixed-term physicians | 0.095**<br>(0.042) |  | 0.047<br>(0.127) | 0.138***<br>(0.048) |  | 0.196<br>(0.157) |
| (6) Population per civil service post (1,000) |  | 0.001<br>(0.005) |  |  | 0.009<br>(0.008) |  |
| *D. Referral (1/0)* |  |  |  |  |  |  |
| (7) Facility hired any fixed-term physicians | -0.060<br>(0.038) |  | -0.005<br>(0.169) | -0.088**<br>(0.039) |  | -0.249**<br>(0.113) |
| (8) Population per civil service post (1,000) |  | 0.000<br>(0.006) |  |  | -0.012**<br>(0.006) |  |

Notes: All regressions control for clinic characteristics (population in catchment area, total number of physicians employed, total value of equipment, whether it is a central township facility, number of village clinics in the township, distance to the nearest county hospital, and net revenue in the year prior to the survey), average values of physician characteristics (years of experience in the facility, medical training, certification level, gender, rural hukou), and disease case fixed effects. F-statistic on excluded instrument (Population in catchment area per civil service post) is 19.0 in Full Sample regressions and 21.2 for Civil Service Only regressions. Robust standard errors shown in parentheses. \*, \*\*, and \*\*\* indicate significance at 10%, 5% and 1%.

## Footnotes

1 See Vegas and De Laat (2003), Bourdon et al. (2010), Atherton and Kingdon (2010), Muralidharan and Sundararaman (2013), Bold et al. (2013), Duflo et al. (2015), Bau and Das (2017; 2020), and Lei et al. (2018).

2 The programs in Kenya and India evaluated by Duflo et al. (2015) and Muralidharan and Sundararaman (2013) gave school committees fixed funds to hire extra contract teachers. As part of their study, Bau and Das (2017; 2020) estimate the effects of a policy change in Pakistan where contract teachers were centrally hired and assigned to schools.

3 Although we don’t find any evidence of spillovers in terms of effort, we do find weak evidence that hiring fixed-term physicians may lead other physicians employed in the same facility to extract more revenue from patients.

4 In China, public facilities (and hence managers) notoriously face incentives to over-treat, however recent reforms, such as the ”Zero Mark-up Policy,” have attempted to address this by de-linking clinic revenues from drug sales.

5 Existing evidence suggests that individuals who select into public service tend to be prosocially motivated, although this depends on the corruption level of the country (Finan et al., 2017). Hanna and Wang (2017) find evidence that college students in India who are more dishonest (as measured in a laboratory experiment) are more likely to prefer a government job.

6 Miller and Babiarz (2014) and Kovacs et al. (2020) review the use of pay-for-performance in developing country health sectors.

7 We also contribute to an inter-related body of work studying factors affecting the performance of health workers (in both the public and private sectors). This literature documents widespread poor quality of care in developing countries and that quality deficits are not only due to a lack of resources or training, but also because front-line healthcare workers also often fail to perform to the best of their ability (Das et al., 2008; Berendes et al., 2011; Kruk et al., 2018; Chaudhury et al., 2006; Leonard and Masatu, 2010b; Mohanan et al., 2015; Sylvia et al., 2017).

8 The quota of personnel at township health centers, the focus of our study, have been around one of every thousand people in the catchment area in the past 40 years (Cao, 1984; SCOPSR, 2011).

9 Within facilities that employ physicians on both types of contracts, the difference in average proportion of pay that is performance based is 8.2 percentage points (p-value *<* 0.01).

10 Tables 1 & 2 report phycian and employment characteristics for the full survey sample. See Appendix Tables 1 & 2 for the sub-sample of physicians for whom performance data were collected. The pattern and magnitude of differences is similar.

11 Definitions for each disease case are shown in Appendix Table 4.

12 Kolmogorov-Smirnov test p-value for distributions in Panel A is 0.096 and 0.036 for Panel B.

13 We use the raw percent of checklist items here rather than an IRT score for comparability between the SP and vignette measures.

14 Analogous regressions that assume linearity (i.e. using the continuous indices for skills and motivation) are presented in Appendix Table 5

15 Oster (2016), presents a method for evaluating omitted variable bias under the assumption that the relationship between treatment status and unobservables can be recovered from the relationship with observables. Following Oster, we use an *R_max_* value of 1.3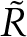, where 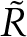 is the r-squared from the full model. In our case 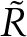 = 0.474, *R_max_* = 0.62.

